# Vitamin D insufficiency and deficiency and mortality from respiratory diseases in a cohort of older adults: potential for limiting the death toll during and beyond the COVID-19 pandemic

**DOI:** 10.1101/2020.06.22.20137299

**Authors:** Hermann Brenner, Bernd Holleczek, Ben Schöttker

## Abstract

**Background:** The COVID-19 pandemic goes along with increased mortality from acute respiratory disease, and measures to limit the spread of the infection go along with increased risk of vitamin D deficiency, especially among high risk groups. It has been suggested that vitamin D_3_ supplementation might help to reduce respiratory disease mortality.

**Methods:** We assessed the prevalence of vitamin D insufficiency and deficiency, defined by 25(OH)D blood levels of 30-50 and <30 nmol/L, respectively, and their association with mortality from respiratory diseases during 15 years of follow-up in a cohort of 9,548 adults aged 50-75 years from Saarland, Germany.

**Results:** Vitamin D insufficiency and deficiency were common (44% and 15%, respectively). Compared to sufficient vitamin D status, respiratory disease mortality was 2.1 (95%-CI 1.3-3.2)- and 3.0 (95%-CI 1.8-5.2)-fold increased, respectively. Although significant increases were seen in both women and men, they were much stronger among women, with 8.5 (95% CI 2.4-30.1) and 2.3 (95% CI 1.1-4.4)-fold increase of respiratory disease mortality in case of vitamin D deficiency among women and men, respectively (p-value for interaction =0.041). Overall, 41% (95% CI 20%-58%) of respiratory disease mortality was statistically attributable to vitamin D insufficiency or deficiency.

**Conclusion:** Vitamin D insufficiency and deficiency are common and account for a large proportion of respiratory disease mortality in older adults, supporting suggestions that vitamin D_3_ supplementation might make a major contribution to limit the burden of the COVID-19 pandemic, particularly among women.

## Introduction

The Corona Virus Disease 2019 (COVID-19) pandemic goes along with strongly increased respiratory disease mortality. It has been suggested that vitamin D_3_ supplementation could be a potentially promising and safe approach to reduce risk of COVID-19 infections and deaths (1). Meta-analyses of randomized clinical trials (RCTs) have shown that vitamin D_3_ supplementation reduces the risk of acute respiratory tract infections (2). Risk reduction with regular (daily or weekly) supplementation of physiological doses of vitamin D was especially strong (by 70%) among people with vitamin D deficiency, but significant risk reduction (by 25%) was also found among people with higher vitamin D levels.

People with pre-existing major diseases, such as diabetes or cancer are at increased risk of dying from severe acute respiratory syndrome coronavirus 2 (SARS-CoV-2) infections. At the same time, prevention of and care for these diseases have been and keep being strongly compromised by current measures to limit the COVID-19 pandemic. Meta-analyses of clinical trials have demonstrated that vitamin D_3_ supplementation has the potential to also reduce cancer mortality by approximately 13% (3).

We previously assessed the prevalence of vitamin D insufficiency and deficiency and their association with all-cause mortality and mortality from cardiovascular, cancer and respiratory diseases in a cohort of 9,548 adults aged 50-75 years from Saarland, Germany (4-8). We aim to present considerably updated and sex-specific follow-up data of 15 years here and to calculate the proportion of respiratory disease mortality that is attributable to vitamin D insufficiency and deficiency. Furthermore, we discuss potential implications for prevention in the context of the ongoing COVID-19 pandemic.

## Methods

### Study design

This investigation is based on the ESTHER study, an ongoing statewide cohort study from Saarland, Germany, details of which have been reported elsewhere (4-9). Briefly, 9,940 men and women, aged 50-75 years at baseline, were recruited by their general practitioners during a routine health check-up between 2000 and 2002. Blood samples were taken at baseline at the general practitioners’ offices. Information on socio-demographic and lifestyle characteristics and medical history were obtained by questionnaires from participants and their general practitioners, and the distribution of those characteristics was similar to the distribution in the respective age categories in the German National Health Survey conducted in a representative sample of the German population in1998 (9). The ESTHER study was approved by the ethics committees of the University of Heidelberg and the state medical board of Saarland, Germany. Written informed consent was issued by all participants.

### Variable assessment

Information on socio-demographic characteristics, lifestyle and diet were obtained by a comprehensive self-administered questionnaire from the study participants at baseline. Height and weight were assessed and documented on a standardized form by the general practitioners during the health check-up. Furthermore, blood and urine samples were taken during the health check-up, centrifuged, sent to the study center and stored at -80°C until analysis.

The most abundant and stable vitamin D metabolite in blood samples, 25-hydroxyvitamin D (25(OH)D) levels was measured from stored serum samples taken at recruitment. The laboratory methods used are described in detail elsewhere (4). In brief, 25(OH)D levels in women were measured with the Diasorin-Liaison analyzer (Diasorin Inc., Stillwater, USA). Analyses in men were conducted in the context of a separate research project several years later (when the Diasorin measurements were no longer offered) with IDS-iSYS (Immunodiagnostic Systems GmbH, Frankfurt Main, Germany). Both immunoassays were standardized retrospectively to the gold standard method liquid chromatography tandem-mass-spectrometry.

Deaths until end of 2016 were identified by inquiry at the residents’ registration offices and death certificates of deceased study participants were provided by local health authorities. The leading cause of death with an ICD-10 code was available for 98.9% of deceased study participants and was coded with ICD-10 codes R98-R99 “unknown cause of death” for 4.4% of deceased participants. These individuals were not excluded and censored at the time of death for cause specific mortality outcomes.

### Statistical methods

Participants of the ESTHER baseline examination (n = 9,940) were excluded from this investigation if no blood sample was available (n = 368) or if they could not be followed-up for mortality (n = 24), which resulted in a total sample size of n = 9,548 subjects for this analysis.

We used the US-American Institute of Medicine’s cut-offs to define adequate vitamin D status (> 50 nmol/L), vitamin D insufficiency (30-50 nmol/L) and vitamin D deficiency (< 30 nmol/L) (10). We assessed prevalences of vitamin D insufficiency and deficiency in the total study population and according to age, sex, life-style factors and major diseases and tested for differences between groups by χ^2^-test.

We compared all-cause, cardiovascular disease, cancer, and respiratory disease mortality between subjects with vitamin D insufficiency or deficiency and subjects with adequate 25(OH)D levels and estimated hazard ratios (HR) with 95% confidence intervals (95%CI) by multivariable Cox proportional hazards models. We used an age, sex and season adjusted model and a full model that was adjusted for potential confounders, which are listed in **Table 1**. Additional adjustment for potential intermediates (cardiovascular disease, history of cancer, diabetes mellitus, hypertension, asthma, total serum cholesterol and serum c-reactive protein levels) did not lead to substantially different results (data not shown). Age and BMI were modelled as continuous variables and all other variables were modelled with the categories shown in **Table 1**. Furthermore, sex-specific analyses were performed and statistical tests on interaction were carried out. In addition, we estimated the population attributable fraction (PAF) of respiratory disease mortality from the prevalences of vitamin D insufficiency and deficiency and their associations with respiratory disease mortality, as derived from our study. The PAF of mortality is the share of mortality in a population that is statistically attributable to a risk factor and that could be avoided by entirely eliminating that risk factor (here: vitamin D insufficiency or deficiency) (11).

**Table 1.** Distribution of characteristics of the study population, and prevalence of vitamin D deficiency and insufficiency by those characteristics

| Characteristic | Proportion of characteristic in total population [%] | Prevalence of vitamin D insufficiency among subjects with characteristic [%] | Prevalence of vitamin D deficiency among subjects with characteristic [%] | p |
| --- | --- | --- | --- | --- |
| Total cohort | 100.0 | 43.8 | 15.1 |  |
| Sex |  |  |  | <0.0001 |
| Female | 56.2 | 52.6 | 15.7 |  |
| Male | 43.8 | 32.6 | 14.2 |  |
| Age (years) |  |  |  | <0.0001 |
| 50-64 | 61.3 | 42.4 | 14.2 |  |
| 65-69 | 22.9 | 45.2 | 15.5 |  |
| 70-75 | 15.8 | 47.3 | 17.7 |  |
| Month of recruitment (Season) |  |  |  | <0.0001 |
| Jan-Feb | 21.7 | 45.7 | 24.4 |  |
| Mar-Apr | 13.9 | 51.6 | 21.2 |  |
| Mai-Jun | 14.2 | 47.8 | 12.4 |  |
| Jul-Aug | 17.0 | 35.6 | 6.9 |  |
| Sep-Oct | 17.0 | 37.0 | 8.1 |  |
| Nov-Dec | 16.2 | 47.0 | 15.6 |  |
| School education |  |  |  | <0.0001 |
| ≤ 9 years | 75.0 | 44.7 | 15.3 |  |
| 9 – 11 years | 14.1 | 44.3 | 13.5 |  |
| ≥ 12 years | 11.0 | 36.1 | 15.4 |  |
| Smoking |  |  |  | <0.0001 |
| Never | 50.5 | 47.9 | 14.5 |  |
| Former | 32.6 | 37.6 | 12.2 |  |
| Current | 16.9 | 43.2 | 21.4 |  |
| BMI (kg/m<sup>2</sup>) |  |  |  | <0.0001 |
| < 30 | 74.5 | 42.1 | 14.1 |  |
| ≥ 30 | 25.5 | 48.9 | 17.9 |  |
| Physical activity <sup>a</sup> |  |  |  | <0.0001 |
| Low | 67.1 | 45.9 | 16.7 |  |
| Moderate or high | 32.9 | 39.6 | 11.6 |  |
| Fish consumption at least once per week |  |  |  | 0.027 |
| No | 33.5 | 44.6 | 15.6 |  |
| Yes | 66.5 | 43.2 | 14.2 |  |

Abbreviations: BMI, body mass index
<sup>a</sup> Defined by ≤ 1 h/week of vigorous physical activity that causes sweating

Missing values for covariates ranged from 0% to 5.8% (for fish consumption). Missing covariate values were imputed with multiple imputation using the Markov Chain Monte Carlo (MCMC) method with 200 burn-in iterations. Twenty data sets were generated. The imputation model consisted of all variables of the full model (modelled as used in the full model) but not the outcome data and the imputation was carried out stratified by sex. All statistical tests were two-sided and the alpha level of significance was set to 0.05.

All statistical analyses were conducted with the software package SAS, version 9.4 (Cary, North Carolina, USA).

## Results

The study population included 43.8% men, mean age was 62.1 years (**Table 1**). Among the 9,548 participants included in the study, 4186 (43.8%) had vitamin D insufficiency (25(OH)D levels of 30 - <50 nmol/L) and 1438 (15.1%) had vitamin D deficiency (25(OH)D levels <30 nmol/L) (**Figure 1A**). **Table 1** also provides a description of further characteristics of the study population at baseline as well as prevalences of vitamin D insufficiency and deficiency according to those characteristics. Both vitamin D insufficiency and deficiency were statistically significantly more frequent among females, with higher age, BMI, in subjects with low physical activity and those who consumed fish less than once per week. A seasonal variation with higher prevalences in winter than in summer months was also observed. Moreover, the prevalence of vitamin D deficiency was significantly higher among current smokers.

**Figure 1.**
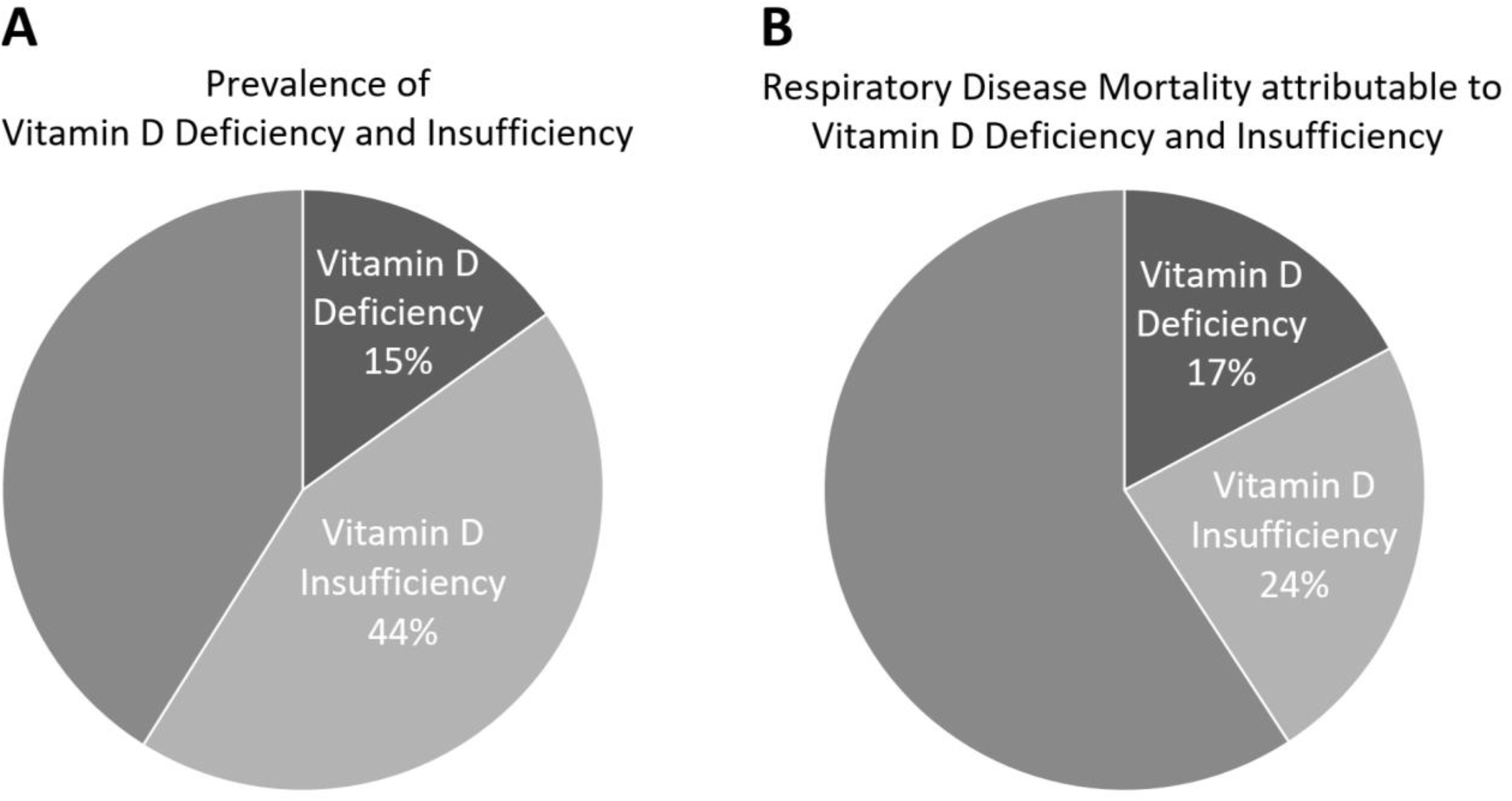
Prevalence of vitamin D deficiency and insufficiency (A) and proportion of deaths from respiratory diseases statistically attributable to these conditions (B)

Overall, 2,363 (24.7%) study participants died during a median of 15.3 years of follow-up and both vitamin D insufficiency and deficiency were associated with significantly increased all-cause mortality compared to sufficient vitamin D status (full model HRs [95%CI]: 1.2 [1.1-1.3] and 1.7 [1.5-1.9], respectively) (**Table 2**). Vitamin D deficiency was also associated with significant increases in CVD and cancer mortality by 52% and 38%, respectively (full model results). However, vitamin D insufficiency and deficiency were particularly strongly associated with respiratory disease mortality with full model HRs of 2.1 (95%CI: 1.3-3.2) and 3.0 (95%CI: 1.8-5.2), respectively. Overall, 41% (95%CI: 20%-58%]) of all deaths from respiratory diseases were statistically attributable to 25(OH)D levels < 50 nmol/L (**Figure 1B**).

**Table 2.**
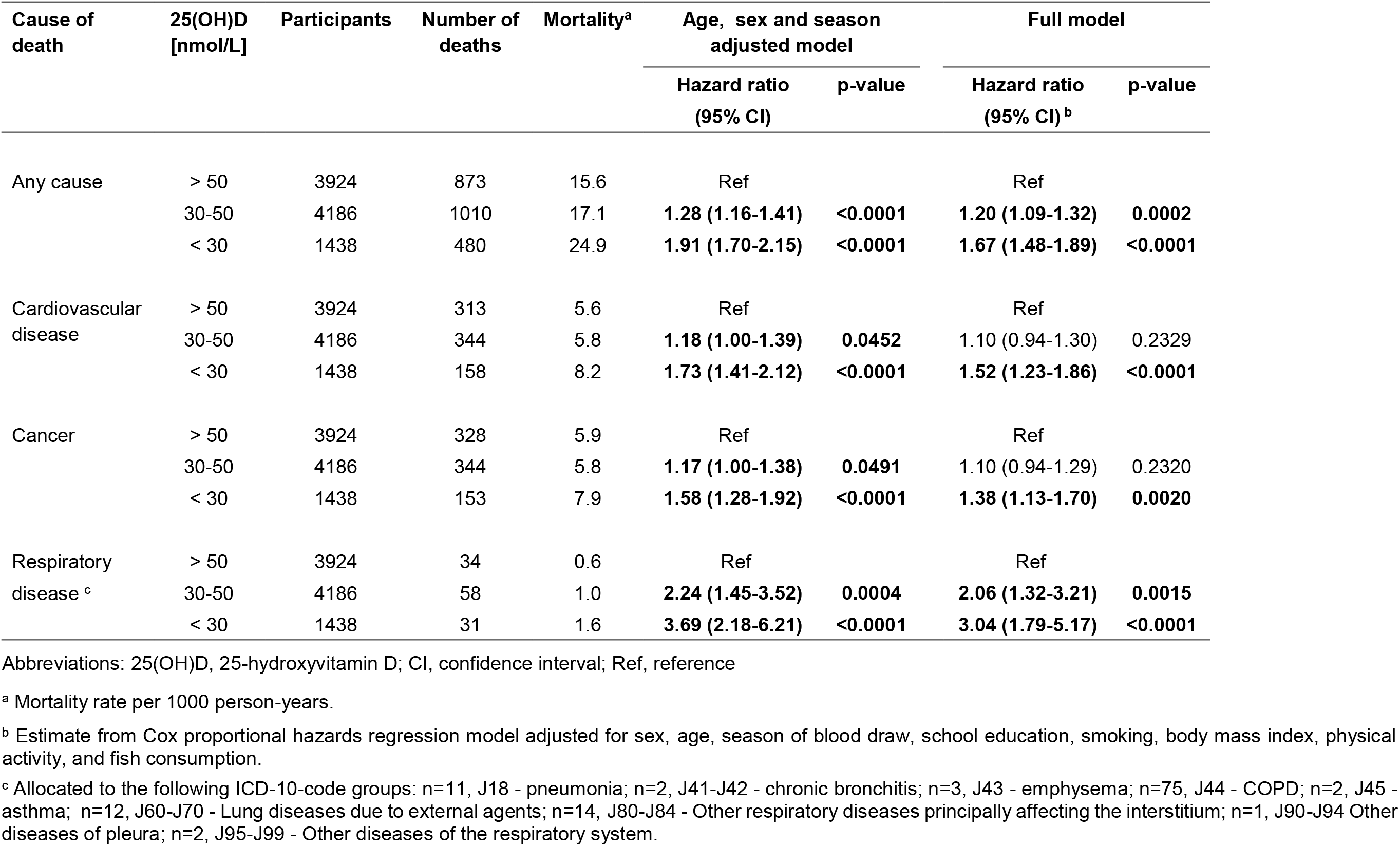
Mortality from major causes of death among people with vitamin D deficiency and insufficiency compared to people with sufficient vitamin D status.

| Cause of death | 25(OH)D [nmol/L] | Participants | Number of deaths | Mortality<sup>a</sup> | Age, sex and season adjusted model |  | Full model |  |
| --- | --- | --- | --- | --- | --- | --- | --- | --- |
|  |  |  |  |  | Hazard ratio (95% CI) | p-value | Hazard ratio (95% CI)<sup>b</sup> | p-value |
| Any cause | > 50 | 3924 | 873 | 15.6 | Ref |  | Ref |  |
|  | 30-50 | 4186 | 1010 | 17.1 | **1.28 (1.16-1.41)** | **<0.0001** | **1.20 (1.09-1.32)** | **0.0002** |
|  | < 30 | 1438 | 480 | 24.9 | **1.91 (1.70-2.15)** | **<0.0001** | **1.67 (1.48-1.89)** | **<0.0001** |
| Cardiovascular disease | > 50 | 3924 | 313 | 5.6 | Ref |  | Ref |  |
|  | 30-50 | 4186 | 344 | 5.8 | **1.18 (1.00-1.39)** | **0.0452** | 1.10 (0.94-1.30) | 0.2329 |
|  | < 30 | 1438 | 158 | 8.2 | **1.73 (1.41-2.12)** | **<0.0001** | **1.52 (1.23-1.86)** | **<0.0001** |
| Cancer | > 50 | 3924 | 328 | 5.9 | Ref |  | Ref |  |
|  | 30-50 | 4186 | 344 | 5.8 | **1.17 (1.00-1.38)** | **0.0491** | 1.10 (0.94-1.29) | 0.2320 |
|  | < 30 | 1438 | 153 | 7.9 | **1.58 (1.28-1.92)** | **<0.0001** | **1.38 (1.13-1.70)** | **0.0020** |
| Respiratory disease<sup>c</sup> | > 50 | 3924 | 34 | 0.6 | Ref |  | Ref |  |
|  | 30-50 | 4186 | 58 | 1.0 | **2.24 (1.45-3.52)** | **0.0004** | **2.06 (1.32-3.21)** | **0.0015** |
|  | < 30 | 1438 | 31 | 1.6 | **3.69 (2.18-6.21)** | **<0.0001** | **3.04 (1.79-5.17)** | **<0.0001** |

Abbreviations: 25(OH)D, 25-hydroxyvitamin D; CI, confidence interval; Ref, reference
<sup>a</sup> Mortality rate per 1000 person-years.
<sup>b</sup> Estimate from Cox proportional hazards regression model adjusted for sex, age, season of blood draw, school education, smoking, body mass index, physical activity, and fish consumption.
<sup>c</sup> Allocated to the following ICD-10-code groups: n=11, J18 - pneumonia; n=2, J41-J42 - chronic bronchitis; n=3, J43 - emphysema; n=75, J44 - COPD; n=2, J45 - asthma; n=12, J60-J70 - Lung diseases due to external agents; n=14, J80-J84 - Other respiratory diseases principally affecting the interstitium; n=1, J90-J94 Other diseases of pleura; n=2, J95-J99 - Other diseases of the respiratory system.

**Table 3** shows the results of the sex-specific analyses. For all-cause, cardiovascular disease and cancer mortality, only modest, statistically non-significant differences were seen between women and men. Although significant increases were seen for respiratory disease mortality in both women and men, they were much stronger among women, with 8.5 (95% CI 2.4-30.1) and 2.3 (95% CI 1.1-4.4)-fold increase of respiratory disease mortality in case of vitamin D deficiency among women and men, respectively (p-value for interaction =0.024).

**Table 3.**
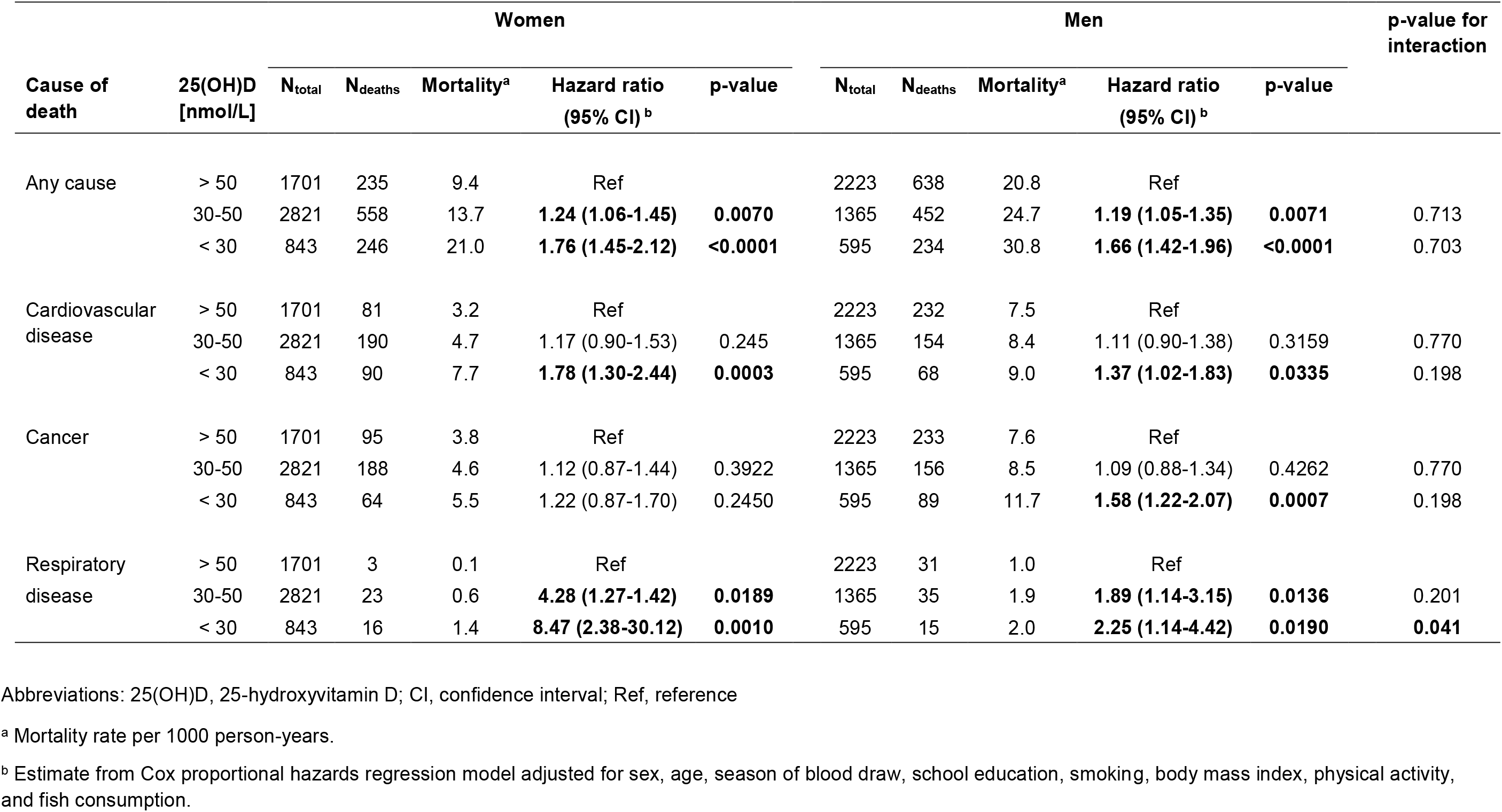
Sex specific analysis on the associations of vitamin D deficiency and insufficiency with mortality from major causes of death.

## Discussion

In this large population-based cohort study from Saarland, Germany, the majority of participants aged 50-75 years at baseline had vitamin D insufficiency or deficiency, and these conditions were associated with increased mortality. In particular, mortality from respiratory diseases was 2.1- and 3.0-fold increased in subjects with vitamin D insufficiency or deficiency, respectively, compared to participants with sufficient vitamin D status. Significant associations with respiratory disease mortality were seen among both women and men, but they were particularly strong for women. Overall, 41% of deaths from respiratory diseases were statistically attributable to vitamin D insufficiency or deficiency, and could possibly be avoided by overcoming these conditions assuming causality of the association. The assumption of causality of vitamin D_3_ effects on mortality obviously requires most careful discussion. Although we made the best attempts to adjust potential confounding factors in our analyses, the major limitation of our study is that we cannot exclude the possibility of residual confounding by imperfect measurement of confounding variables, such as smoking or physical activity, or omission of unknown confounders in this observational study. As addressed in detail elsewhere (6), interpretation of the evidence is further complicated by the fact that vitamin D deficiency could be considered both a consequence of poor health as well as a risk factor for increased vulnerability to acute disease and poor outcomes of chronic diseases among people with poor health. It is therefore paramount to critically evaluate our findings in the light of additional criteria and evidence, such as biological mechanisms and plausibility, and, in particular, in the light of data from RCTs providing vitamin D_3_ supplementation.

Deaths from respiratory disease are mostly deaths from exacerbations of COPD which are caused by acute infections in the majority of cases or from acute respiratory infections causing pneumonia. Vitamin D_3_ is thought to protect from occurrence and poor outcomes of respiratory infections by several mechanisms, including enhanced physical barriers (maintenance of junction integrity), cellular innate immunity, and adaptive immunity (1). Innate and adaptive immunity are being influenced by sex hormones (12), which may explain the observed interaction of sex and 25(OH)D levels with respiratory disease mortality. According to data from the US-American National Health and Nutrition Survey women have a higher inflammation burden than men (the age-range was 40-90+ years (13)). Especially postmenopausal women, like those included in the ESTHER study, have a high inflammatory burden because a decline in estrogen level during menopause is associated with an increased expression of pro-inflammatory cytokines, incl. interleukin 6 and tumor necrosis factor (TNF) α (12,14,15). A cytokine storm as an adverse immune response to a SARS-CoV-2 infection is currently a major hypothesis for the underlying cause of a large proportion of COVID-19 deaths (16). Sufficient 25(OH)D levels are suggested to contribute to prevention of the cytokine storm (17, 18). This may be especially important in postmenopausal women who tend to have both lower 25(OH)D levels and a higher inflammatory burden than men.

Although specific data for the role of vitamin D_3_ in protecting from SARS-CoV-2 infections and their consequences are yet to be published it appears plausible to assume that these mechanisms would be relevant for this infection in a similar manner as for other severe viral respiratory diseases, such as influenza. In a meta-analysis of individual participant data of 25 RCTs that included 11,321 participants, aged 0–95 years, vitamin D_3_ supplementation was shown to reduce the risk of acute respiratory tract infection (OR 0.88, 95% CI 0.81–0.96) (2). The best effects were shown for daily or weekly vitamin D_3_ supplementation without additional bolus doses (OR 0.81, 95% CI 0.72 to 0.91). The protective effects were particularly strong in those with baseline 25-hydroxyvitamin D levels <25 nmol/L (OR 0.30, 95% CI 0.17– 0.53), suggesting an approximately 3-fold risk among people with vitamin D deficiency not receiving vitamin D_3_ supplementation compared to people with vitamin D deficiency receiving supplementation. In a recent meta-analysis of RCTs on vitamin D_3_ supplementation for patients with COPD, the risk of acute exacerbations was estimated to be reduced by 61% (95% CI 36%-77%) (19). In remarkable consistency with our results, these meta-analyses of RCTs provide strong evidence for the preventive potential of vitamin D_3_ supplementation against acute respiratory infections and COPD exacerbations in particular.

A first study posted on a pre-print server on May 13, 2020 suggests that the protective effects of vitamin D_3_ on other acute respiratory tract infections may be translated to COVID-19 infections (20). Vitamin D deficiency and vitamin D_3_ treatment data were available for 499 COVID-19 patients from Chicago for the year prior COVID-19 testing. Being likely vitamin D deficient (defined as being vitamin D deficient at last available time point without increase of vitamin D treatment) at the time of COVID-19 testing was associated with a 1.8-fold increased risk of being tested positive for COVID-19 (p<0.02) as compared to likely vitamin D sufficient.

It is worth noting that beneficial effects of vitamin D_3_ supplementation against manifestation or exacerbation of acute respiratory infection during an epidemic would be expected to go beyond individual protection of those using supplementation, as limiting such manifestation and exacerbation would also be expected to reduce the potential of spread of the disease to other persons and relieve the overload of the medical system by the epidemic.

To our knowledge, no previous vitamin D_3_ supplementation RCTs have addressed mortality from respiratory disease as primary endpoint, and no meta-analysis of results for this specific endpoint have been reported, which most likely reflects the relatively small share of deaths from this endpoint among all deaths. In our cohort of older adults these deaths accounted for 5.2% of all deaths. Even though this proportion is expected to be higher during the COVID-19 pandemic, the majority of deaths still occur from other diseases, and the summary effect, benefit-harm ratio and cost-effectiveness with respect to all relevant outcomes therefore deserve most careful attention for any general prevention efforts.

In that respect, vitamin D_3_ supplementation appears to be a particularly promising approach, especially for population groups with high prevalence of vitamin D insufficiency or deficiency, such as the elderly and those with severe comorbidities (which essentially coincide with population groups at highest risk of severe course and death from SARS-CoV-2 infection (21)): The personal, health care and societal costs of the vitamin D_3_ intervention are negligibly low compared to the very high costs of currently employed “general population measures”, such as extensive testing for the infection and the lockdown of large proportions of economic and social life, including the delay or omission of much of routine medical care for other relevant diseases. In fact, some of these measures are expected to severely aggravate vitamin D insufficiency or deficiency, especially in high risk groups, such as restrictions of spending time outdoors for the total population (as practiced, for example, in France and Spain) or certain high risk groups, such as nursing home residents (as practiced in many countries including Germany). Such restrictions dramatically reduce opportunities to maintain adequate vitamin D levels through endogenous synthesis by relevant sun exposure.

Vitamin D_3_ supplementation has been demonstrated to be safe in numerous large scale studies, and the risk of harm seems to be negligible compared to the risk of harmful side effects of the aforementioned and other general population measures, such as delayed diagnosis and treatment of cancer, myocardial infarction or stroke, withheld or deferred delivery of surgical or other medical services, or health risks related to unemployment and loneliness (22-25). On the contrary, one expected “side effect” would be reducing total cancer mortality by 13%, as suggested by a recent meta-analysis of RCTs (3). For Germany, with currently approximately 230,000 deaths from cancer per year (26), this would translate in prevention of approximately 30,000 cancer deaths each year, suggesting substantial additional benefit besides lowering the COVID-19 burden during the COVID-19 pandemic and beyond.

In conclusion, our results, along with evidence from meta-analyses from RCTs regarding results of vitamin D_3_ supplementation on various outcomes, suggest that vitamin D_3_ supplementation could make a major contribution to lowering mortality from respiratory and other diseases during and beyond the COVID-19 pandemic, in particular among women. The Endocrine Society recommends 1,500-2,000 IU vitamin D_3_/day for adults of any age at high risk for vitamin D deficiency (27). The costs for such supplementation are in the order of 30 € per person per year, or even half that amount when sufficient vitamin D supply is ensured by carefully dosed sun exposure during the summer months. Along with expected savings from prevented respiratory and other diseases, this would make vitamin D_3_ supplementation a particularly cost-effective and most likely cost-saving measure, whose currently still widely neglected potential should receive increased attention and should more widely be utilized in the fight against the COVID-19 pandemic and beyond.

## Data Availability

The ESTHER data cannot be made publicly available due to legal restrictions. However, data can be shared on the basis of research proposals that are in accordance with the study's aims.

## Acknowledgements

The ESTHER study was funded by grants from the Saarland state Ministry for Social Affairs, Health, Women and Family Affairs (Saarbrücken, Germany), the Baden-Württemberg state Ministry of Science, Research and Arts (Stuttgart, Germany), the Federal Ministry of Education and Research (Berlin, Germany) and the Federal Ministry of Family Affairs, Senior Citizens, Women and Youth (Berlin, Germany). The authors have no conflicts of interest to disclose. The author contributions were as follows: HB designed the research and drafted the manuscript, BS conducted statistical analyses, and BH critically read and commented the manuscript and added aspects to the discussion. All authors contributed to the data collection for this project.

